# Deep Phenotyping of Blood Cell Data Reveals Novel Clinical Biomarkers

**DOI:** 10.64898/2026.03.24.26349221

**Authors:** Ya-Lin Chen, Cindy Zhang, Fabienne Lucas, Jennifer Hadlock, Brody H Foy

## Abstract

**Introduction:** The complete blood count with differential (CBD) is one of the most commonly performed blood tests worldwide, used in nearly all areas of medicine. Although modern CBD analyzers generate flow-cytometry based single-cell measurements, the resultant CBD markers are limited to coarse summary features, such as total cell counts and average cell sizes. This means, the markers cannotdetect subtle cell population shifts that may signal early-stage pathogenesis. To test this, we evaluate whether AI-based analysis of the raw single-cell data underlying the CBD can be used to develop novel, clinically prognostic biomarkers, across patient settings.

**Method:** We developed two complementary methods for biomarker discovery using CBD tests and evaluated them with longitudinal data from an academic medical center. To create interpretable biomarkers, we clustered cells into physiologically meaningful sub-populations and performed robust statistical summarization. In tandem, self-supervised autoencoders were developed to extract novel non-linear markers. We evaluated the utility of these clustering (CLS) and autoencoder (AE) markers for patient prognostication across a range of outcomes (mortality, inpatient admission, and future disease development).

**Results:** Our study included 242,623 CBD samples from 127,545 patients. Both clustering and embedding approaches successfully generated hundreds of new clinical biomarkers. Many biomarkers showed strong prognostic associations for all-cause mortality, inpatient admission, and development of anemia, cancer, or cardiovascular disease, with associations remaining significant after adjustment for demographics and clinical CBD markers. A large subset of these prognostic markers also showed high novelty – having low correlations to existing CBD markers, while also exhibiting significant correlations with broader physiologic signals, such as inflammatory, hormonal, infectious, and coagulopathic markers.

**Conclusion:** Collectively, these results demonstrate how modern AI techniques can allow for deeper phenotyping of routine clinical blood counts, generating novel biomarkers that capture more subtle physiologic signals than what are currently clinically utilized.

## Introduction

The complete blood count and white cell differential (CBD) – one of the most frequently collected clinical tests worldwide^1^ – is primarily based on single-cell light scatter measurements, collected through basic flow cytometry techniques^2^. Although the process of cell-type annotation differs from traditional multi-parameter flow cytometry, where cell populations are typically defined by manual gating, analyzer-based cytometers rely on the same flow cytometry principles to measure cellular properties and use automated algorithms to classify cell sub-populations. This means that alongside standard CBD markers such as red cell (RBC), red cell distribution width (RDW), platelet (PLT), and white cell counts (WBC), analyzers typically generate rich ancillary data streams. These include cell population data (CPD) markers^3^: additional cell markers created by the device manufacturers; as well as the raw single-cell data: multi-channel, single-cell measurements of tens of thousands of cells. Despite this data richness, clinically reported CBD results remain largely restricted to a set of count parameters and mean properties, leaving most single-cell information unexploited.

Most CPD markers – such as cell size and activation levels – are classified as “research parameters”, typically excluded from clinical reporting despite their potential value^4^. However, recently the FDA granted 510(k) clearance for monocyte distribution width (MDW), a CPD measure of monocyte size heterogeneity, for sepsis risk assessment^5^, highlighting the clinical value of these data streams.Beyond this translational success, many research studies provide evidence of diagnostic value of CPD markers across a range of conditions, such as infectious diseases^6,7^, hematologic disorders^8–10^, and in general prognostic settings^11^.

However, while CPD markers have potential clinical value – they are hindered by lack of technical clarity, with their construction and reporting typically made at the discretion of each device manufacturer, often with only limited documentation. This means potentially informative single-cell signals cannot be systematically evaluated. Despite their being many established analytical and computational methods for analysis of single-cell data^12,13^, prior CBD-focused efforts have only involved the use of manual gating to isolate cell sub-types^14,15^, and no existing work has methodically approached systematic, unbiased generation of novel CBD-derived markers. Given the clinical ubiquity of the CBD, this is a major gap.

Here, we developed two complementary approaches to deep phenotyping of CBD data, to generate novel biomarkers. We used unsupervised learning to identify cell populations and computed statistical summaries to characterize each population, providing a dense set of interpretable markers. In tandem, we trained a series of self-supervised autoencoders to extract compact biomarkers which capture non-linear interactions across the cell populations. Through analysis of multiple major health conditions, we show that these deep phenotypic markers are highly novel, provide significantly enhanced diagnostic and prognostic signals, and provide insights into broader physiologic signals.

## Methods

### Study design

We designed our study to systematically extract novel markers from raw CBD single-cell data for deeper hematologic phenotyping (**Figure 1**). We employed two complementary approaches, each addressing distinct limitations of conventional CBD and CPD marker generation. First, we used clustering methods to identify major cell sub-types within each flow channel, then computed statistical summaries of cellular features within each cell population. This approach preserved biological interpretability while capturing distributional properties beyond simple means and cell counts. Second, we trained self-supervised autoencoder models to derive compact, non-linear representations from the single-cell data without relying on predefined cell types. Together, these methods provided biologically grounded features and non-linear features that extend beyond conventional CBD and CPD markers. We then evaluated these newly derived markers across a variety of diagnostic and prognostic scenarios by assessing their associations with health outcomes and non-CBD tests.

**Figure 1.**
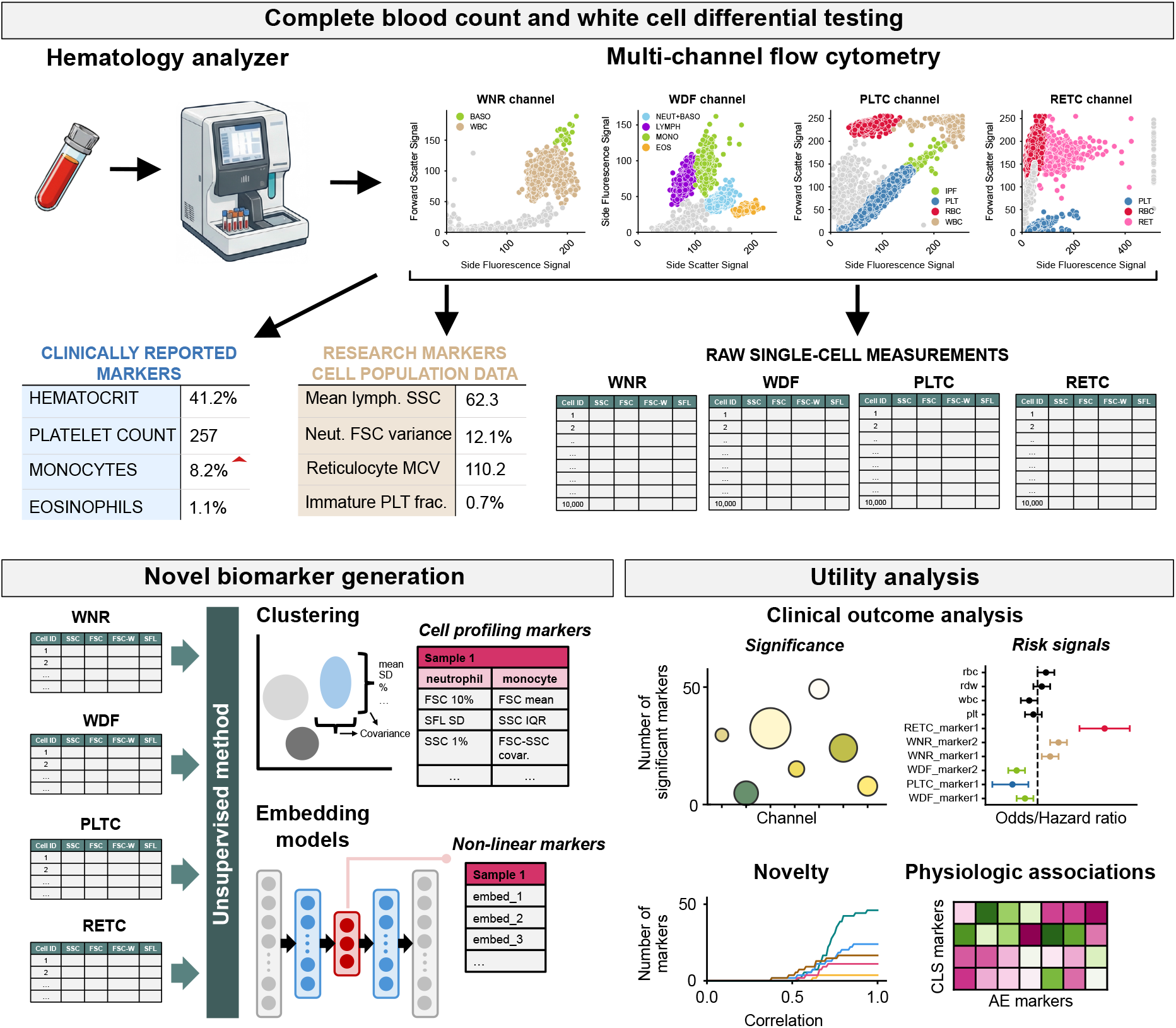
Study workflow for AI-derived biomarker discovery. Routine complete blood count (CBC) tests are processed by hematology analyzers using flow cytometry, in up to four modes: WNR for total white cell counts (WBC) and nucleated cell counts; WDF for white cell differential counts; PLTC for specializing platelet testing; and RETC for red cell and reticulocyte testing. For each mode, the analyzer produces three data types: clinically reported markers (e.g., CBC and differential counts); “research markers” – referred to as cell population data (CPD) markers – that are not available in clinical reports; and the raw single-cell flow cytometry data. We extracted these raw flow cytometry files, and generated novel biomarkers using two approaches: unsupervised clustering with statistical summarization and self-supervised autoencoder embeddings to capture non-linear sample representations. We evaluated the resulting biomarkers through utility analyses, including associations with major clinical outcomes, assessments of novelty based on correlation with CBC, differential counts, and CPD markers, and correlations with broader non-CBC laboratory tests. Blood tube and hematology analyzer icons were generated by Gemini 3 and are intended to be illustrative only.

### Cohort and flow cytometry data

We collected flow cytometry files (.fcs file format) derived from all adult (18-100yr old) CBD tests run across outpatient (OP), inpatient (IP), and emergency room (ER) settings at the University of Washington Medical Center (UWMC) between 01-April 2024 and 27-August 2025. All tests were performed on Sysmex XN-1000 analyzers, which produced three data types: clinically reported CBD markers, research CPD markers (defined in **Supplementary Table S1**), and raw single-cell data matrices. Each fcs file included one or more channels – corresponding to different types of analysis performed by the analyzer, focused on: white cells and basophils (WNR, available for 99.0% of samples), the white cell differential (WDF, 61.3%), platelets (PLTC, 7.7%), and red cells and reticulocytes (RETC, 2.0%). Within each channel, cells were characterized by four scatter features: side scatter (SSC, correlated with cellular complexity and/or granularity), forward scatter (FSC, correlated with cell size), forward scatter width (FSC-W, measuring cell aggregation or doublets), and side fluorescence (SFL, correlated with cellular activation including nucleic acid content)^16^. Cell types within each channel could be distinguished by their characteristic positions in two-dimensional scattergrams (**Supplementary Figure S1**).

For each patient encounter, we selected the first available CBD test, retained the most recent single-cell file per channel (for samples that were run multiple times), and excluded samples with missing channel-specific cell types (**Supplementary Table S2**). The first year of data (Apr-2024 – Mar-2025) was used as a training set for the clustering and autoencoders. Data from Apr-2025-Aug-2025 comprised the test set for evaluating model performance and outcome associations. A random 20% of the training set was used as a validation set for hyper-parameter tuning. All patients from the training set were excluded from the test set. To allow for computational tractability, autoencoder models were trained with a random 10,000 samples from the training set, for the WNR and WDF channels. Scatter values for each flow measurement were normalized by dividing by the standard upper limit for the measurement (511 for SFL in the RETC channel, 255 for all other measurements and channels).

### Interpretable biomarker generation

To generate novel biomarkers from the single-cell data, we pursued two approaches: generation of detailed cell features through clustering and summarization (referred to as CLS markers), and generation of non-linear features through autoencoder embedding models (AE markers). For the CLS markers we first generated clusters of known cell types (RBC, lymphocytes, eosinophils, etc.) from the scattergrams. Then, we generated biomarkers by summarizing the cellular characteristics of each cluster. Clustering was performed using FlowSOM, which applies self-organizing maps (SOM) followed by consensus hierarchical clustering^17^. We first initialized a grid of nodes with random weights, iteratively updating them based on individual cell marker expression profiles. We optimized the approach with batched SOM^18^, which processed multiple samples simultaneously, with incremental training^19^, to update the model across successive batches for a maximum of 30 iterations (see **Supplementary Methods**). The resulting SOM nodes were then grouped using consensus hierarchical clustering to identify distinct cell populations. For each channel, two scatter features were selected based on their scattergrams and the number of clusters was determined a priori according to the channel-specific cell types (**Supplementary Table S2**)^2^. To evaluate clustering performance, we computed the correlation between analyzer-generated and clustering-generated cell counts and percentages.

Following clustering, for each cell type (neutrophils, RBC, etc.), we derived the following statistical features for each measurement channel: the 1st, 5th, 10th, 25th, 50th (median), 75th, 90th, 95th and 99th percentiles, the mean, standard deviation, and minimum and maximum value. We also considered the pairwise covariances among the four scatter features, yielding 58 features per cell type. In total, there were 116 WNR channel markers, 290 WDF markers, 174 PLTC markers, and 174 RETC markers.

### Non-interpretable biomarker generation

A primary limitation of clustering and summarization is the inability to capture features arising from highly non-linear interactions between cells or across cell types. To address this, we trained three autoencoder models to generate compact, non-linear biomarkers: a feedforward network (FF)^20^, convolutional neural network (CNN)^21^, and a transformer-based model^22^ (referred to as the set model, see **Supplementary Methods** for detailed model descriptions). The FF autoencoder used single-layer fully connected encoder and decoder on flattened cell data. The CNN autoencoder, inspired by Deep CNN^23^, applied a convolutional layer (kernel size 1 × 4 markers) followed by average pooling across cells, then reconstructed input via transposed convolution. Both FF and CNN models generated embeddings without modeling cell-cell interactions. In contrast, the transformer autoencoder implemented the Fully Differentiable Set Autoencoder (FDSA) framework^22^, which used recurrent neurons with attention mechanisms to treat the single-cell data as unordered sets and explicitly model cell-cell interactions. We evaluated nine configurations for each autoencoder type by varying the number of input cells per sample (256, 512, or 1024) and the size of the embedding layer (16, 64, or 128), with each configuration trained separately for each analyzer channel. To account for the variability of number of cells per sample, each sample was either down- or up-sampled with replacement to reach the desired size. While training, we assessed model performance every two epochs using validation loss, computed as mean squared error (MSE) for the FF and CNN models and as MSE plus cross-entropy for the set model. Training was halted when validation loss failed to decrease by more than 1% for five consecutive evaluations (10 epochs). We used an initial learning rate of 0.001 that decreased by 20% every 5 epochs^24^, a batch size of 256 samples, and the Adam optimizer^25^.

### Outcome association analysis

We evaluated associations of CBD, CPD, CLS, and AE markers with five major clinical care setting and outcomes: these included inpatient admission within 48hrs for ER patients; 30-day all-cause post-discharge mortality for inpatients; and future diagnosis of iron deficiency anemia, cancer, and major adverse cardiovascular events (MACE; myocardial infarction, heart failure, stroke) for outpatients. All diagnoses were defined by International Classification of Disease (ICD) codes, listed in **Supplementary Table S3**. Outcome data for IP admission and mortality were collected on 30-September 2025, with deaths pulled from health record data, linked to the US Social Security Death Index^26^. We excluded patients in the ER cohort if they were admitted within 48hrs of the study end date (30-Sep-2025); and in the IP cohort if admitted within 30 days of the study end date. For the OP cohort, final diagnoses were collected on 16-October 2025. We excluded patients with any prior history of the diagnoses above, as well as those attaining these diagnoses within 7 days after the CBD test (as these reflect cases that were likely known or suspected at time of CBD testing). All OP patients were censored based on the date of their last known interaction with the UW medical system, up until 16-October 2025 (the last data pull date), and patients censored within 7 days after their CBD test were excluded.

IP admission and mortality associations were calculated using logistic regression, reported as odds ratios (ORs). Disease associations were measured through time-to-event analysis, via Cox proportional hazards models, reported as hazard ratios (HRs). All associations were adjusted by age, sex, four canonical complete blood count (CBC) indices (RDW, RBC, WBC, PLT) and five white cell differential (DIFF) markers (neutrophil, basophil, eosinophil, monocyte, lymphocyte percentage).Unless otherwise noted, all ORs and HRs were normalized to a 1 standard deviated change in the marker.

### Correlation analysis

To analyze associations of AE markers with broader physiologic signals, we collected all quantitative non-CBD laboratory tests ordered concurrently to our study’s CBD samples. We then computed Spearman correlations between each marker and these non-CBD tests, for all tests with at least 50 samples. To identify novel correlations with non-CBD laboratory tests, we defined novel AE markers as those whose maximum absolute correlation with CBD or CPD markers was below 0.5.. For completeness, key results are presented across additional correlation thresholds. Results for correlations with non-CPD tests were reported for all AE markers with at least one moderate absolute correlation (above 0.3).

### Statistical analysis

Statistical significance was defined as a two-sided p-value < 0.05 unless otherwise noted. Significance was based on t-tests for continuous outcomes, and chi-squared tests for event rates. Clustering was performed in R 4.3.1, and all other analyses were performed in Python. The set autoencoder was run with Python 3.7, brcpytorch v0.1.3, and PyTorch v1.13.1; all other analyses were performed with Python 3.9.

## Results

An overview of the study design is given in **Figure 1**. The test dataset encompassed 40,927 samples, with characteristics summarized in **Table 1**. Samples with PLTC testing (N=2,551) represented an older (mean 58.05 [SD 17.56]) and predominantly male population (54.17%), obtained largely from IP settings (48.06%), and with the highest cancer prevalence across channels (1.39%). Conversely, RETC tests were mainly ordered in OP settings (51.80%) and exhibited the lowest median RBC (3.60 IQR [2.85-4.23]), the highest median RDW (14.60 IQR [13.20-16.90]), and the highest rate of outcomes except for cancer.

**Table 1.** Cohort characteristics.

|  | WNR | WDF | PLTC | RETC |
| --- | --- | --- | --- | --- |
| Test set N | 36,929 | 24,810 | 2,551 | 1,058 |
| Age, mean (SD), years | 50.62 (18.43) | 50.35 (18.40) | 58.05 (17.56) | 55.77 (18.33) |
| Sex, Male % | 41.57 | 41.46 | 54.17 | 50.47 |
| CBD test location, % |  |  |  |  |
| ER | 15.54 | 21.6 | 15.05 | 8.88 |
| IP | 13.9 | 12.13 | 48.06 | 39.32 |
| OP | 70.57 | 66.27 | 36.89 | 51.8 |
| Complete blood count markers, median (IQR) |  |  |  |  |
| RBC, 10<sup>6</sup>/uL | 4.52<br>(4.12-4.90) | 4.52<br>(4.11-4.90) | 3.97<br>(3.16-4.57) | 3.60<br>(2.85-4.23) |
| RDW, % | 13.00<br>(12.40-13.90) | 13.10<br>(12.40-14.00) | 14.10<br>(13.00-15.90) | 14.60<br>(13.20-16.90) |
| PLT, 10<sup>3</sup>/uL | 246.00<br>(202.00-295.00) | 246.00<br>(202.00-297.00) | 125.00<br>(96.00-150.00) | 211.00<br>(147.00-282.00) |
| WBC, 10<sup>3</sup>/uL | 6.98<br>(5.58-8.90) | 6.96<br>(5.53 - 8.92) | 6.76<br>(4.94-9.60) | 6.83<br>(4.88-9.37) |
| Outcomes, N (%) |  |  |  |  |
| IP admission | 5737 (28.08) | 5360 (28.60) | 384 (45.31) | 94 (78.72) |
| Mortality | 5132 (4.35) | 3009 (6.58) | 1226 (10.28) | 416 (16.35) |
| Anemia | 22969 (1.58) | 14395 (1.77) | 727 (3.44) | 240 (9.17) |
| Cancer | 23232 (0.77) | 14583 (0.60) | 722 (1.39) | 471 (1.27) |
| MACE | 24680 (0.48) | 15509 (0.57) | 817 (0.61) | 479 (1.04) |

Abbreviations: CBD, complete blood count with differentials; ER, emergency room; IP, inpatient; OP, outpatient; RBC: red blood cells; RDW, red cell distribution width; PLT, platelet; WBC, white blood cell; Anemia, iron deficiency anemia; MACE, major adverse cardiovascular events.

### Clustering recapitulates known cell types

Unsupervised clustering derived cell groups that showed strong agreement with CBD cell counts (**Figure 2a-c**), suggesting it accurately recapitulated known cell types. Samples with cell counts in the interquartile range showed higher correlation than samples with counts outside that range (**Figure 2b**). Overall, predicted cell counts demonstrated a correlation of at least 0.9 with channel-reported markers, except for reticulocytes (**Figure 2c**). This discrepancy likely stemmed from the continuous maturation of reticulocytes into red blood cells, leading to a less clearly defined cell population boundary.

**Figure 2.**
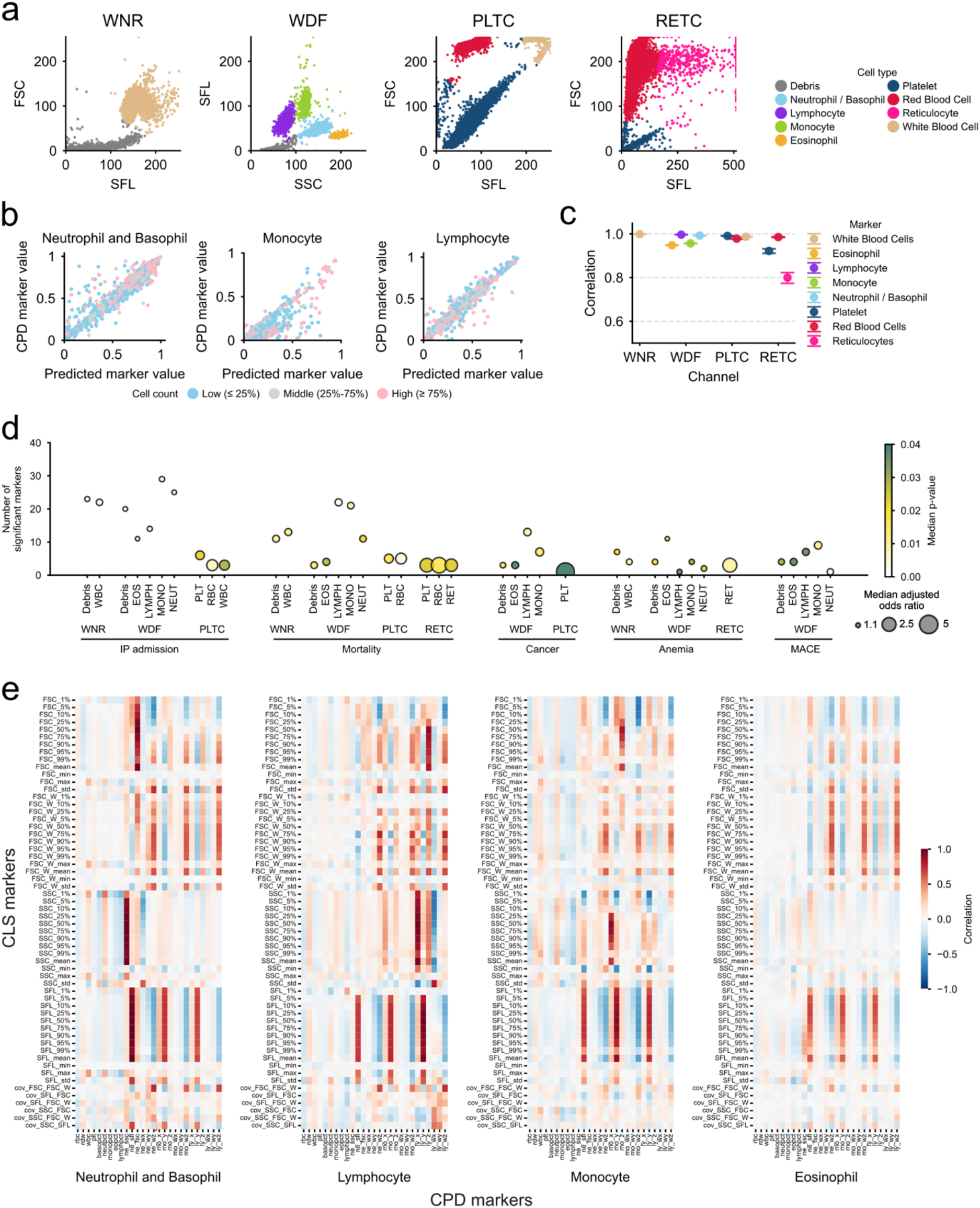
Clustering identified novel markers associated with health outcomes. **a**. Scattergrams of the clustering results of four CBC analyzer channels. **b**. Correlation between predicted and machine-generated cell-type measures in WDF channel, stratified by the number of cells per sample. **c**. Correlation between predicted and machine-generated cell-type measures across four flow cytometry channels. **d**.Number of clustering-derived markers significantly associated with clinical outcomes, stratified by cell-type and flow channels. **e**. Correlation between CPD markers and clustering markers in WDF channel. Odds ratios in (**d, e**) were adjusted for age, sex, four major CBC markers (platelets, red cells, red cell distribution width, and white cells) and five DIFF markers (neutrophil, basophil, lymphocyte, and eosinophil percentages). All odds ratio were normalized to the risk increase per 1std increase in the marker. Abbreviations for (**d**) include EOS, eosinophil; LYMPH, lymphocytes; MONO, monocytes; NEUT, neutrophils; RET, reticulocytes. Other abbreviations are defined in the methods.

### Cluster-derived markers exhibit strong prognostic signals

Following clustering, CLS markers were defined through robust characterization of cluster statistical properties (measurement percentiles, correlations, variance, etc.). Many CLS markers were significantly associated with major clinical outcomes after adjustment for demographics and CBD values (**Figure 2d**), with strong signals seen across all outcomes and all channels. IP admission exhibited the strongest overall signals, especially for markers related to the WNR and WDF channels – with particular prominence among monocyte (N=29) and neutrophil (N=25) population markers. For mortality, RETC RBC markers (RBC, median OR=1.775, median p=0.003) showed the highest ORs whereas WDF lymphocytes markers (N=22, p<0.001) were most numerous. Across diagnostic outcomes, the WDF channel produced the largest array of significant risk markers, with contributions across all major cell types.

Unsurprisingly, many CLS markers demonstrated high correlations with CPD markers (**Figure 2e**). Covariance between scatter features showed low correlations with CPD markers (median absolute correlation=0.105), and many of these derived features remained associated with clinical outcomes after adjustment for CBD markers, suggesting they provided complementary information to existing markers. Interestingly, some CLS markers showed strong correlations across cell types. For example, neutrophil SFL markers correlated with CPD markers of non-neutrophil SFL features, suggestive of coregulation across blood cell populations^27^.

### Set autoencoders generate clinically relevant embeddings

To complement clustering, we trained three autoencoder models (FF, CNN, set) to capture non-linear representations of the single-cell data. Model training time did not vary consistently with input or embedding size (**Figure 3a**); however, set autoencoders required significantly more epochs to train than the FF or CNN models (mean [SD] 60.9 [19.4] vs. 54.4 [18.6] and 24.0 [3.6] respectively) (**Figure 3b**). Their more complex structure allowed set models to achieve the lowest reconstruction errors, which was especially noticeable for bimodally distributed features (**Figure 3c**). Across input and embedding sizes, the set models achieved lower MSE loss than the FF and CNN models (mean [SD] 0.003 [0.002] vs. 0.017 [0.006] and 0.014 [0.0001] respectively, p<0.001) (**Figure 3d**). Overall, set models outperformed FF and CNN models in reconstruction by achieving lower model loss and more efficient embeddings.

**Figure 3.**
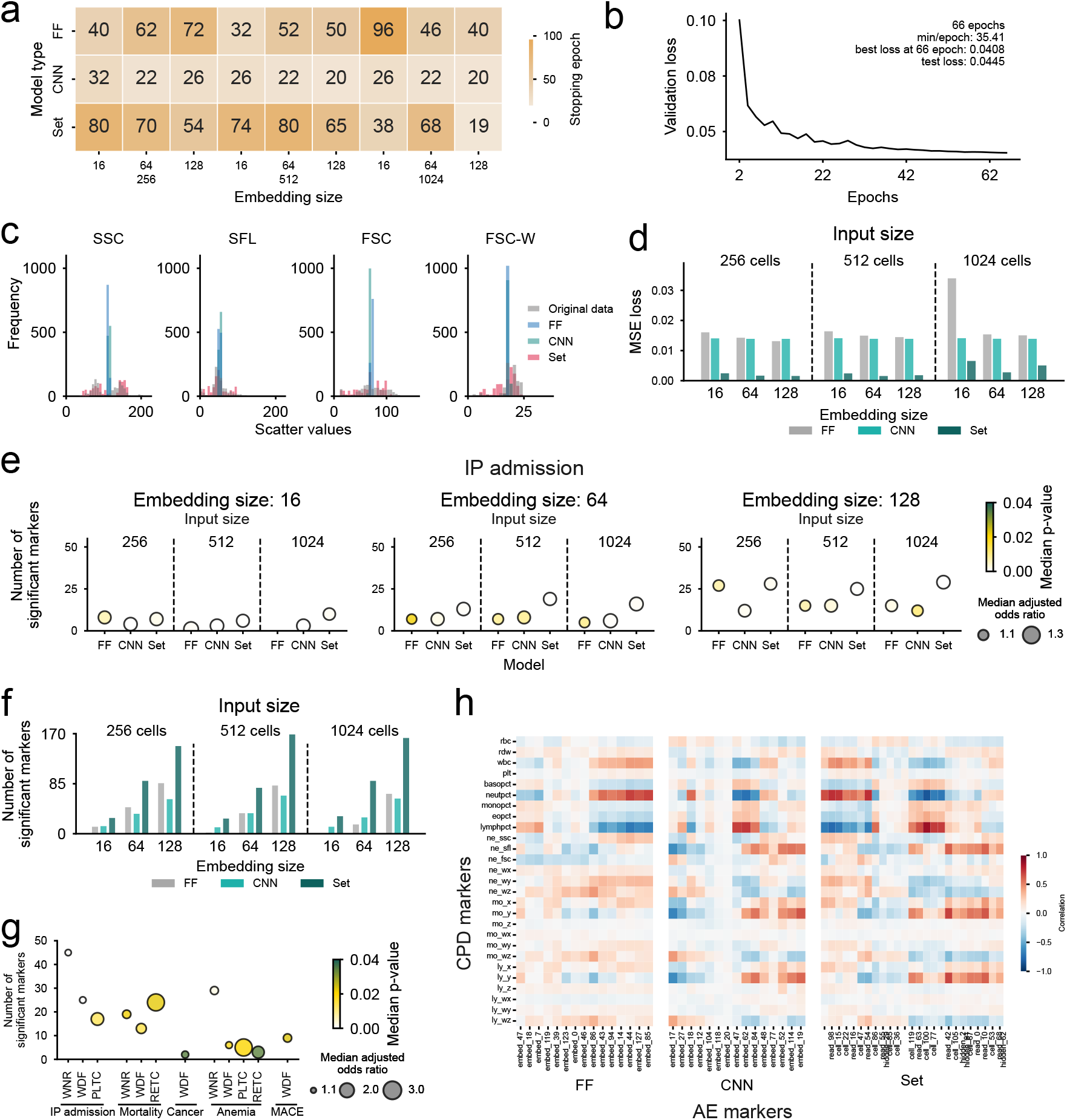
Set autoencoders outperformed FF and CNN models and derived clinically relevant markers. **a**. Number of training epochs required for convergence across autoencoder architectures and configurations. b. Learning loss curve of the set model with 512 input cells, and 128-size embedding, showing validation loss across training epochs. **c**. Reconstruction outputs compared to the input cell distribution from the feed-forward (FF), convolution (CNN), and set autoencoders, with 1024 input cells and a 128-size embedding layer. **d**. Test MSE loss for each autoencoder type and configuration. **e**. Number of significant markers for IP admission from WDF channel. **f**. Total number of significant markers across all clinical outcomes. Counts are summed across four independently trained, channel-specific models for each model configuration. **g**. Marker significance across channel and outcomes from set models of 512 input cells and 128-size embedding layer. **h**. Correlation between CPD markers and autoencoder-derived markers trained with 512 input cells and 128-size embedding layer that are significantly associated with IP admission. Odds ratios in (**e, g**) were adjusted for age, sex, four major CBC markers (platelets, red cells, red cell distribution width, and white cells) and five DIFF markers (neutrophil, basophil, lymphocyte, and eosinophil percentages). All odds ratio were normalized to the risk increase per 1std increase in the marker. Abbreviations are defined in the methods. Markers in **f** that had significant associations with multiple outcomes were only counted once.

### Autoencoder-derived markers capture diverse prognostic risks

Across ER, OP and IP settings, many AE markers were strongly associated with major clinical outcomes. AE markers with higher-dimensional embeddings produced more statistically significant associations, and set models yielded the largest number of significant markers (**Figure 3e-f**). Set models also produced more informative embeddings that revealed small, distinct patient subgroups with markedly worse clinical status that were not detected by FF or CNN embeddings (**Supplementary Figure 2**). Given this, we chose to focus remaining analysis on the set model with a 512 cell input size and a 128 embedding layer, which produced 169 novel clinical markers. The largest set of novel markers was derived for IP admission, using the WNR channel (N=45, median OR=1.14, p<0.001), which also produced the largest array of significant markers for anemia (**Figure 3g**). Conversely, the most significant markers for mortality, cancer, and MACE were produced by the WDF channel. Correlation patterns between AE markers and existing markers (CBD and CPD) showed how non-interpretable embeddings captured many of the dominant hematologic phenotypes of the CBD and CPD markers (**Figure 3h**). In particular, FF markers correlated strongly with cell counts – suggesting they captured fewer complex features than the other embedding models, which additionally correlated with more functional cell features (e.g., mean neutrophil side fluorescence, etc.).

### Flow cytometry derived markers provide highly novel clinical risk signatures

To assess novelty of the CLS and AE markers, we computed their pairwise correlations with CBD and CPD markers (**Figure 4a**). Unsurprisingly, the overall set of CLS and AE markers were strongly correlated with existing CBD and CPD signals, with AE markers showing higher correlations (median max. absolute correlation with CBC=0.32, DIFF=0.19, CPD=0.65) than CLS markers (median max. absolute correlation with CBC=0.23, DIFF=0.12, CPD=0.60). After adjustment for age, sex, and CBD markers, many CLS and AE markers remained significantly associated with major clinical outcomes, often equaling or outperforming the magnitude of risk stratification achieved by canonical CBD risk markers (RBC, PLT, WBC, RDW) (**Figure 4b-c**). Considering the AE markers (**Figure 4b**), the strongest associations were among markers from WNR and WDF channels – partly due to the largest cohort sizes (as specialized PLTC and RETC analysis are far less commonly performed, **Table 1**).

**Figure 4.**
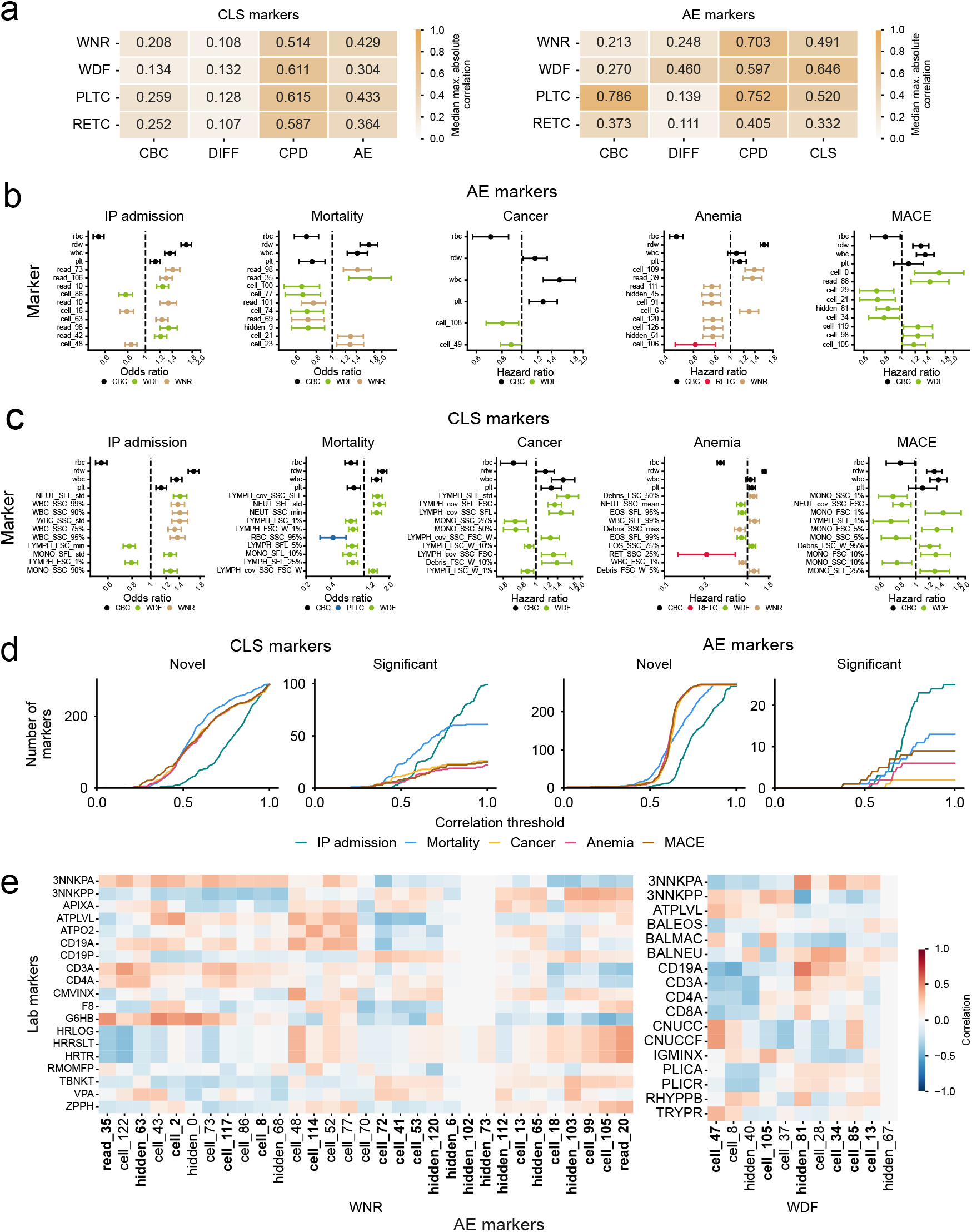
AI-derived markers capture phenotypes beyond CBD and CPD markers. **a**. Median of the maximum correlations of AE and CLS markers with CBC, DIFF, CPD markers, and each other. **b-c**. The 10 most significant AI-derived markers (ranked by p-value), compared to four CBC (RBC, RDW, WBC, and PLT), for each major outcome. (**b** of AE markers, **c** of CLS markers). **d**. Cumulative number of novel and outcome-significant markers with correlations below a given threshold to CBD and CPD markers. **e**. Correlation between AE markers (512 input cells and 128-size embedding layer) and non-CBD markers. Odds ratios in (**b, c**) were adjusted for age, sex, four major CBC markers (platelets, red cells, red cell distribution width, and white cells) and five DIFF markers (neutrophil, basophil, lymphocyte, and eosinophil percentages). All odds ratio were normalized to the risk increase per 1std increase in the marker. Abbreviations of non-CBD markers are defined in **Supplementary Table S4** and in the methods.

Considering CLS markers, increased cell variance or shifts in the tails of properties were the strongest risk markers for most outcomes (**Figure 4c**). For example, variance in neutrophil side fluorescence (NEUT_SFL_std) was strongly associated with both mortality (OR=1.53 [1.30-1.80], p<0.001) and IP admission (OR=1.42 [1.31-1.54], p<0.001), and the low tail of monocyte forward scatter (MONO_FSC_1%, a correlate of the size of the smallest monocytes) was strongly associated with future MACE risk (HR=1.47 [1.14-1.89], p=0.002). The major exception was future cancer risk, which was driven more strongly by covariance of lymphocyte properties (e.g., covariance between lymphocyte SSC and SFL [LYMPH_cov_SSC_SFL], HR=1.48[1.22–1.78], p<0.001), and mean monocyte properties (e.g., median monocyte SSC [MONO_SSC_50%], HR=0.73 [0.60–0.88], p=0.001). This supports the interpretation that signatures of high clinical relevance in the raw data are in the tails of the cell distributions – which are not captured by commonly used CPD markers.

To assess clinical novelty of both the CLS and AE markers, we next determined how many markers retained significant associations with clinical outcomes (after adjustment for age, sex, and CBD markers) while concurrently having low correlation to all existing CBD and CPD markers (**Figure 4d**). We found that both the CLS and AE approaches produced novel markers (maximum absolute correlation with CBD/CPD markers < 0.5), while still having significant clinical associations. This suggests that deep phenotyping of the CBD data can produce general prognostic risk markers that provide substantial information beyond what is currently available.

### Novel autoencoder markers capture broader physiologic signals

The primary advantage of the AE markers – i.e., they capture non-linear data features and cell-cell interactions – comes with a trade-off in greatly reduced interpretability. However, we hypothesized that this non-linearity may allow AE markers to capture broader physiologic signals. To assess this, we measured AE marker correlations with all concurrently measured laboratory tests across our cohorts (**Figure 4e**). For both the WNR and WDF channels, many AE markers (including those which were both novel and significant from **Figure 4b**) showed moderate-to-good correlations with broader laboratory tests (max absolute correlation: 0.521). Many AE markers were associated with more specific blood cell properties, such as white cell subset counts^28^, including CD4+ (cluster of differentiation) cells (CD4A, max absolute correlation=0.402), and blood cell counts from bronchoalveolar lavage (BAL) testing (e.g., BAL macrophages [BALMAC], max absolute correlation=0.357). AE markers also showed significant correlations with infectious disease tests, such as cytomegalovirus (CMVINX, max absolute correlation=0.403) and human immunodeficiency viruses (HRLOG, max absolute correlation=0.422). Furthermore, we identified associations with specific homeostatic markers or testing, including coagulation-related indices, such as factor VIII activity (F8, max absolute correlation=0.381) and apixaban level (APIXA, corr: 0.280), in addition to anemia-related markers, such as glucose-6-phosphate dehydrogenase test (G6HB, max absolute correlation=0.483), an assay for an enzyme deficiency causing hemolytic anemia, and zinc-protoporphyrin/heme ratio (ZPPH; max absolute correlation=0.321), a marker for pre-anemic iron deficiency diagnosis. Further descriptions of each of these tests is given in **Supplementary Table S4**.

## Discussion

Here, we developed a multi-pronged approach to generate novel biomarkers from routinely collected CBD testing. CBD-derived markers provided strong prognostic signals across many different patient outcomes and collecting settings, independent of existing CBD marker signals. A subset of novel markers also showed strong connections to broader physiologic signals, such as detailed blood cell sub-types, infection, and inflammation signals.

To our knowledge, no prior group has used CBD flow-cytometry data at scale to derive novel clinical biomarkers. In recent years, multiple groups have investigated potential added value of CBD flow cytometry data for specific target conditions, such as infectious diseases^6,7^, hematologic disorders^8–10^, and perioperative risk^29^; however, these approaches relied on analyzer-generated markers, limiting their ability to produce systematic profiles. Recently, a few groups have sought to explore the direct use of raw flow cytometry and/or single-cell data for end-to-end outcome prediction^14,15,30^, such as through multi-modal modelling of scattergrams alongside CBD markers for leukemia prediction^30^ – with similar efforts occurring for other non-CBD data sources^23,31,32^.

Comparative to these approaches, we believe our methodology has multiple significant advantages. Firstly, the focus on biomarker generation allows for enhanced clinical relevance, as biomarkers can be more easily integrated into holistic clinical care in standard human-in-the-loop frameworks, comparative to more advanced black-box algorithms. This is of particular relevance to the blood count, which is already used in complex, diverse, and multifactorial settings across medicine^33^. Secondly, our framework for CLS marker generation allows for clear clinical interpretability. Flow cytometry measurements correlate strongly with major physiologic signals, such as cell size or activation levels, meaning cell distribution characteristics can be cleanly interpreted^16^. For example, increased neutrophil forward scatter variance (ne_fsc) corresponds to increase variance in neutrophil size, analogous to the widely used red cell distribution width – a canonical general risk marker^34^. This is further supported by the recent FDA approval of monocyte distribution width as a prognostic marker for sepsis^5^. As such, our work allows for generation of novel, interpretable biomarkers from single-cell data, which provides a framework for targeted investigation of the utility of specific biomarkers for specific clinical decisions. While our study focused on the CBD test, collection of cell-level quantitative measurements occurs across many other clinical and biological settings (e.g., immunologic flow cytometry, single-cell omics, etc.). Our study provides a proof of concept that statistical and machine learning analysis of these data streams can enable the creation of novel clinical biomarkers. Of particular note, the strongest risk signatures we identified were primarily due to measures of cell variance, or measurements of extreme cell tails. This is logical, as most mean or central cell characteristics would already be well described by existing CBD markers, such as RDW of mean red cell volume (MCV). It may also suggest that many physiologic shifts in mean cell characteristics are likely preceded by shifts in distributional tails. This aligns with prior findings from our group, such as evidence of increasing red cell microcytosis preceding broader mean red cell volume decline^35^. This finding may be of particular relevance for MACE, where variance and tail markers exhibited larger hazard ratios than standard CBD markers (**Figure 3**), consistent with prior evidence that shifts in white cell differential counts are predictive of cardiovascular diseases^36^.

Results in **Figure 4** showed that the derived markers provided novel information, compared to existing CBD and CPD markers. Interestingly, CLS markers demonstrated greater novelty than AE markers, despite likely capturing more linear cell features and interactions. This may be explained by the fact that autoencoders are optimized to minimize reconstruction loss, which favors extracting dominant structural information, such as cluster size distributions, aligning with reported behavior of the set autoencoder model^37^. Despite this, many AE markers showed significant correlations with broader laboratory tests, including non-hematologic assays. Correlations with white cell subsets and immunoglobulin assays suggest that AE markers could potentially enhance clinical test targeting, informing who may benefit from additional work-up. This is of particular relevance given the low-cost, rapid nature of CBD testing, compared to many resource-intensive tests which may require substantial turnaround times or may be non-viable in resource-limited healthcare settings.

In summary, the analysis of raw flow cytometry data from the complete blood count generated both interpretable and non-linear biomarkers. Extracted markers captured unique physiologic signatures, while providing novel prognostic signals for both general outcomes and specific disease states. Given flow cytometry is widely used across medicine, our approach can substantially increase information extraction from both routine and targeted clinical tests.

## Declarations

### Ethics approval and consent to participate

The study was performed under a research protocol approved by the University of Washington Medicine Institutional Review Board, under a waiver of informed consent due to minimal risk.

### Data availability

Due to IRB restrictions on sharing of protected health information, individual patient data cannot be shared.

### Code availability

Clustering analysis was performed using the FlowSOM R package (v.2.10.0; https://www.bioconductor.org/packages/release/bioc/html/FlowSOM.html). Set autoencoder analysis was implemented using the FDSA package (https://github.com/PaccMann/fdsa) Other analysis was performed using Python version 3.9.

### Author contributions

BF and YC were responsible for study design. Data collection and analysis was performed by YC with support from CZ. BF, JH and FL contributed to study supervision. All authors contributed to data interpretation, and writing, and have read and approved the final manuscript.

## Acknowledgements

The authors thank Nathan Breit, Patrick Mathias, and the UW DLMP Informatics team for assistance with extraction and archiving of the CBD, CPD, and single-cell data.

